# COVID-19 Epidemic Outside China: 34 Founders and Exponential Growth

**DOI:** 10.1101/2020.03.01.20029819

**Authors:** Yi Li, Meng Liang, Xianhong Yin, Xiaoyu Liu, Meng Hao, Zixin Hu, Yi Wang, Li Jin

## Abstract

**Background:** In December 2019, pneumonia infected with a novel coronavirus burst in Wuhan, China. Now the situation is almost controlled in China but is worse outside China. We aimed to build a mathematical model to capture the global trend of epidemics outside China.

**Methods:** In this retrospective, outside-China diagnosis number reported from Jan 21 to Feb 28, 2020 was downloaded from WHO website. We develop a simple regression model on these numbers:

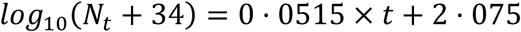

where N_t_ is the total diagnosed patient till the *i*th day, t=1 at Feb 1.

**Findings:** Based on this model, we estimate that there have been about 34 unobserved founder patients at the beginning of spread outside China. The global trend is approximately exponential, with the rate of 10 folds every 19 days.

**Research in context:** *Evidence before this study:* In December 2019, pneumonia infected with a novel coronavirus burst in Wuhan, China. Now the situation is almost controlled in China but is worse outside China. Now there are 4,691 patients across 51 countries and territories outside China. We searched PubMed and the China National Knowledge Infrastructure database for articles published up to Feb 28, 2020, using the keywords “COVID”, “novel coronavirus”, “2019-nCoV” or “2019 novel coronavirus”. No published work about the global trend of epidemics outside China could be identified.

*Added value of this study:* We built a simple “log-plus” linear model to capture the global trend of epidemics outside China. We estimate that there have been about 34 unobserved founder patients at the beginning of spread outside China. The global trend is approximately exponential, with the rate of 10 folds every 19 days.

*Implications of all the available evidence:* With the limited number of data points and the complexity of the real situation, a straightforward model is expected to work better. Our model suggests that the COVID-19 disease follows an approximate exponential growth model stably at the very beginning. We predict that the number of confirmed patients outside China will increase ten folds in every 19 days without strong intervention by applying our model. Powerful actions on public health should be taken to combat this epidemic all over the world.

## Introduction

In early December of 2019, pneumonia cases of unknown cause emerged in Wuhan, the capital of Hubei province, China.^1^ A novel coronavirus (now named as COVID-19) was verified and identified as the seventh member of enveloped RNA coronavirus (subgenus sarbecovirus, Orthocoronavirinae subfamily) through high-throughput sequencing.^2-4^ Human-to-human transmission in hospital and family settings had been accumulating _5-7_ and has occurred among close contacts since the middle of December 2019 based on evidence from early transmission dynamics.^8^ According to WHO statistics, the accumulating number of diagnosed patients in China was 78,961 in Feb 28, 2020.^9^

COVID-19 raised intension not only within China but internationally. Since the first case of pneumonia caused by COVID-19 was reported from Wuhan, COVID-19 patients had rapidly been diagnosed in other cities of China and the neighboring countries including Thailand, South Korea, Japan, and even to a few Western countries.^10-12^ On January13 2020, the Ministry of Public Health of Thailand reported the first imported case of lab-confirmed novel coronavirus (2019-nCoV). ^13^ As of Feb28 2020, cases had been reported in 52 countries and territories. The number of confirmed COVID-19 cases had reached 4,691 outside China, ^9^ and the number is still increasing. South Korea, the worst affected country outside China, has reported another jump. The 571 new cases brought South Korea’s total confirmed cases to 2,337. A surge in cases of COVID-19 in Italy, Japan and Iran also heightened fears that the world is on the brink of a pandemic.^14^ In Feb 28, WHO increased the assessment of the risk of spread and risk of impact of COVID-19 to very high at the global level.

Recently, many researchers devoted to more details about the analysis of the spread of epidemic.^14,15^ Some paralleled results have shown that the reproductive number R0 estimated of COVID-19 is bigger than that of SARS based on different models. ^16-18^ With the limited number of data points and the complexity of the real situation, a straightforward model is expected to work better (see Discussion). In this study, we propose a “log-plus” model for the situation prediction which only requires daily number of total diagnosis outside China. This model assumes that there were some unobserved founder patients at the beginning of spread outside China and exponential growth later. Despite the simplicity of our model, it fits the data well (R^2^=0.991). The prediction is expected to provide practical significance on social application and evidence for enhancing public health interventions to avoid severe outbreaks.

## Methods

### Data

Daily diagnosis number of countries outside China is download from WHO situation reports (https://www.who.int/emergencies/diseases/novel-coronavirus-2019/situation-reports). The data in this analysis starts from Jan 21 and currently it ends at Feb 28.

### Model

We first explored the data by plotting log transformed daily number. We observed a good linear trend recently, but a relatively poor fit at the beginning of spread. We therefore consider that there are some unobserved founder patients at the beginning. Based on exploratory data analysis and mathematical intuition we proposed the following model:

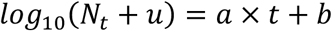

where N_t_ is the number of diagnosed patients outside China according to WHO at the *t*th day, t=1 at Feb 1; u is the number of unobserved founder patients at the beginning of spread outside China; a and b are simple linear regression parameters. We enumerated u from 0 to 100 with step size 1. For each u, we calculated Pearson’s correlation R^2^ between t and *log*_10_ (*N*_*t*_ +*u*). We selected the *û* that maximize R^2^ and estimate corresponding *â* and 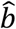 using a simple linear regression between t and *log*_10_ (*N*_*t*_ +*û*).

### Availability of source code

The source code of model estimation is at: https://github.com/wangyi-fudan/COVID-19_Global_Model

## Results

### Data Table

The WHO daily count of diagnosis number outside China and its “log-plus” transform as well as model fit were shown in table 1.

**Table 1:** WHO Daily Diagnosis Number Outside China.

| Date | t | $N_t$ | $\log_{10}(N_t+34)$ | model fit |
| --- | --- | --- | --- | --- |
| 21-Jan | -10 | 4 | 1.580 | 1.559 |
| 22-Jan | -9 | 4 | 1.580 | 1.611 |
| 23-Jan | -8 | 7 | 1.613 | 1.662 |
| 24-Jan | -7 | 11 | 1.653 | 1.714 |
| 25-Jan | -6 | 23 | 1.756 | 1.765 |
| 26-Jan | -5 | 29 | 1.799 | 1.817 |
| 27-Jan | -4 | 37 | 1.851 | 1.868 |
| 28-Jan | -3 | 56 | 1.954 | 1.920 |
| 29-Jan | -2 | 68 | 2.009 | 1.971 |
| 30-Jan | -1 | 82 | 2.064 | 2.023 |
| 31-Jan | 0 | 106 | 2.146 | 2.075 |
| 1-Feb | 1 | 132 | 2.220 | 2.126 |
| 2-Feb | 2 | 146 | 2.255 | 2.178 |
| 3-Feb | 3 | 153 | 2.272 | 2.229 |
| 4-Feb | 4 | 159 | 2.286 | 2.281 |
| 5-Feb | 5 | 191 | 2.352 | 2.332 |
| 6-Feb | 6 | 216 | 2.398 | 2.384 |
| 7-Feb | 7 | 270 | 2.483 | 2.435 |
| 8-Feb | 8 | 288 | 2.508 | 2.487 |
| 9-Feb | 9 | 307 | 2.533 | 2.538 |
| 10-Feb | 10 | 319 | 2.548 | 2.590 |
| 11-Feb | 11 | 395 | 2.632 | 2.641 |
| 12-Feb | 12 | 441 | 2.677 | 2.693 |
| 13-Feb | 13 | 447 | 2.682 | 2.744 |
| 14-Feb | 14 | 505 | 2.732 | 2.796 |
| 15-Feb | 15 | 526 | 2.748 | 2.847 |
| 16-Feb | 16 | 683 | 2·856 | 2·899 |
| 17-Feb | 17 | 794 | 2·918 | 2·950 |
| 18-Feb | 18 | 804 | 2·923 | 3·002 |
| 19-Feb | 19 | 924 | 2·981 | 3·053 |
| 20-Feb | 20 | 1,073 | 3·044 | 3·105 |
| 21-Feb | 21 | 1,200 | 3·091 | 3·156 |
| 22-Feb | 22 | 1,402 | 3·157 | 3·208 |
| 23-Feb | 23 | 1,769 | 3·256 | 3·259 |
| 24-Feb | 24 | 2,069 | 3·323 | 3·311 |
| 25-Feb | 25 | 2,459 | 3·397 | 3·362 |
| 26-Feb | 26 | 2,918 | 3·470 | 3·414 |
| 27-Feb | 27 | 3,664 | 3·568 | 3·466 |
| 28-Feb | 28 | 4,691 | 3·674 | 3·517 |

### Parameter Estimation

*û, â* and 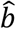 were estimated as 34, 0·0515 and 2·075 respectively according to Feb 28 data (figure 1)

**Figure 1:**
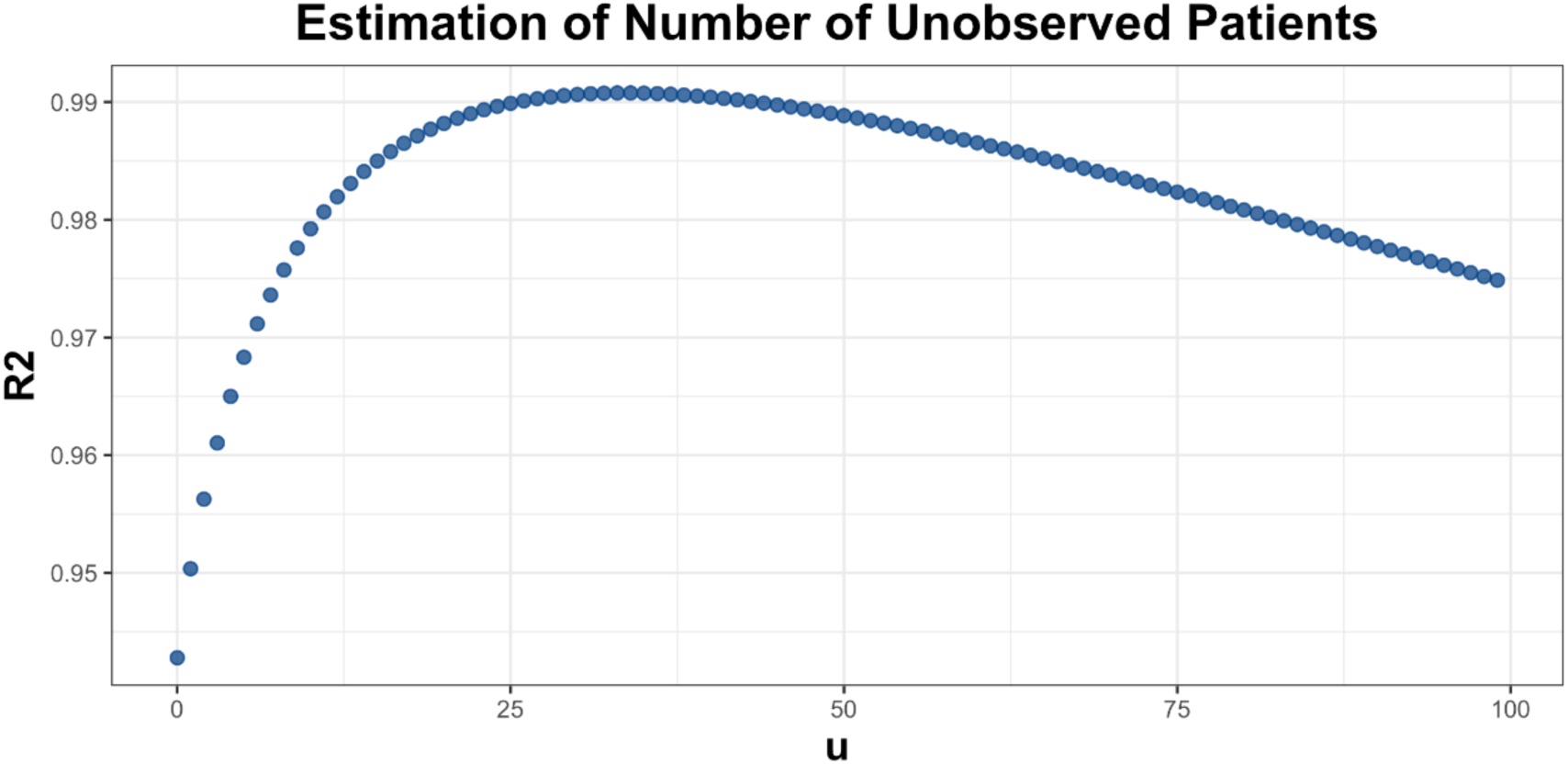
Estimation of u Parameter by Enumeration.

### Global Trend Model

We plot *log*_10_ (*N*_*t*_ + 34) against time to visualize the model fitting (figure 2). The model R^2^ is 0·991 indicating an excellent model fit.

**Figure 2:**
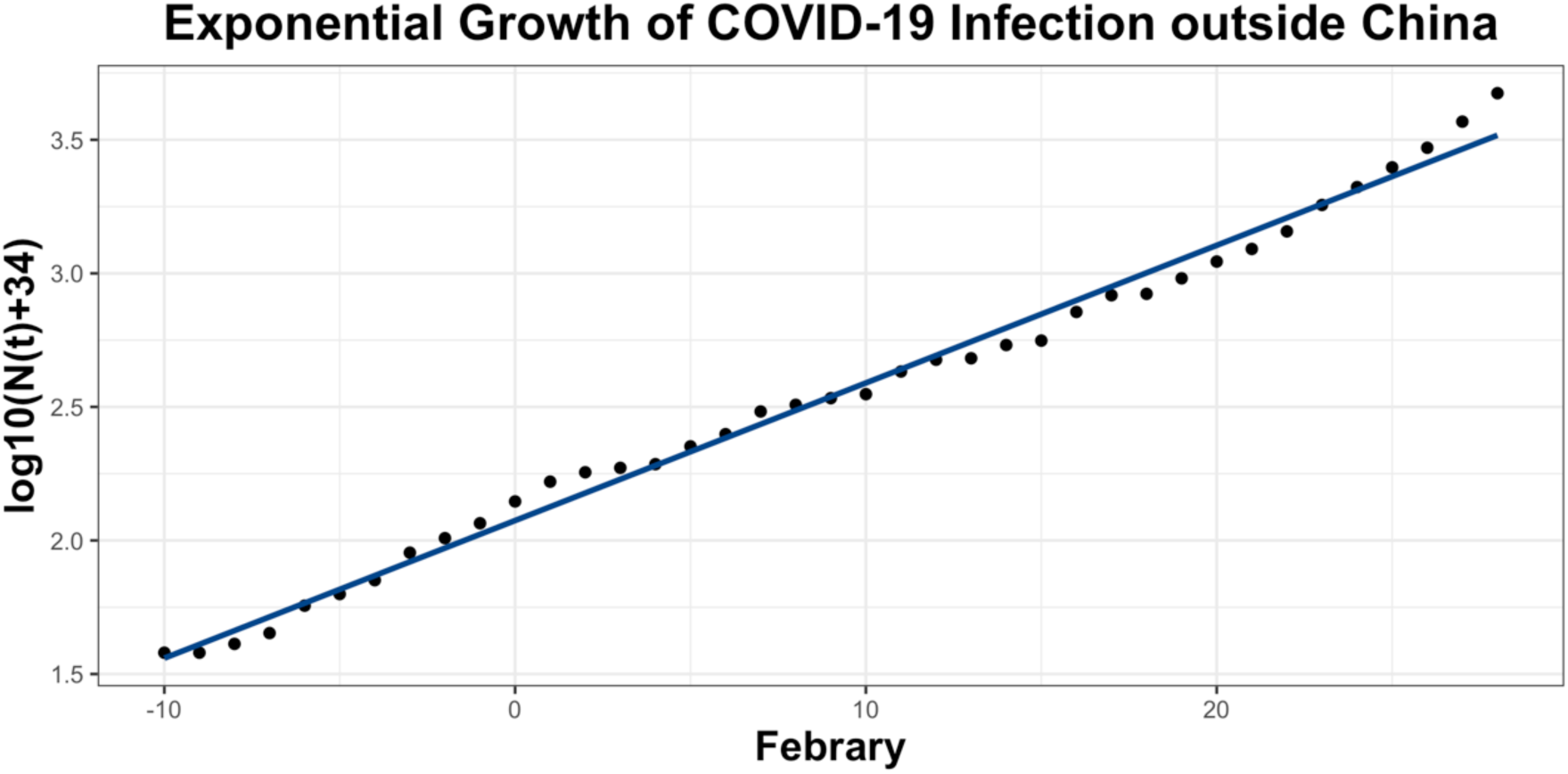
The Exponential Growth of COVID-19 Infection Outside China.

### Future Number Prediction

Currently the number of diagnoses at Febuary 28 is 4,691. We predict that the number of diagnoses outside China will expand exponentially with the rate of 10 folds every 19 days without strong intervention.

### Conclusion

Based on this model, we estimated that there have been about 34 unobserved founder patients at the beginning of spread outside China. Our model suggests that the COVID-19 disease follows an approximate exponential growth model stably at the very beginning. The situation is dangerous as we expect 10 folds increases of patients every 19 days without strong intervention. We call on world-wide strong actions on public health referring to experiences learnt from China and Singapore.

## Discussion

In this report, we only analyzed the total number of diagnoses outside China. The country scale data is available but is not as complete as total number. Here, we limited this analysis to capture the global trend.

This model is a minimal extension of the ‘default’ exponential growth model with the estimation of 34 unobserved founder patients outside China. An almost perfect model fitting (R^2^=0·991) indicates that the spread of disease does follow our model.

A simple and straightforward linear model inherits with some advantages: (1) it works on small sample size due to limited observation or somewhat unperfect data. (2) it is relatively robust in complex situations. The virus spreading pattern is complex and yet varies across the world. A simple model simplified the situation and provides coarse-grained trend estimation. (3) A linear model has nice extrapolation ability comparing to complex models (e.g. Neural network).

The existent of 34 unobserved founder patients is not surprising. They may have mild symptoms thus did not went to the hospitals. However, we did not preclude the possibility that they already existed before or parallel with Wuhan’s burst.

## Data Availability

Daily diagnosis number of countries outside China is download from WHO situation reports (https://www.who.int/emergencies/diseases/novel-coronavirus-2019/situation-reports).

## Contributors

YW conceived the idea and wrote the source code. YW, YL, ML and LJ contributed data analysis, generating tables and figures, and manuscript writing. YL, ML, XY, LX, MH, ZH, YW, LJ contributed the theoretical analysis and manuscript revision. All authors contributed to final revision of the manuscript.

## Acknowledgments

We thank the Fudan University High-End Computing Center for supporting computations involved in this study.

